# Individual bioenergetic capacity as a potential source of resilience to Alzheimer’s disease

**DOI:** 10.1101/2024.01.23.23297820

**Authors:** Matthias Arnold, Mustafa Buyukozkan, P. Murali Doraiswamy, Kwangsik Nho, Tong Wu, Vilmundur Gudnason, Lenore J. Launer, Rui Wang-Sattler, Jerzy Adamski, The Alzheimer’s Disease Neuroimaging Initiative, Alzheimer’s Disease Metabolomics Consortium, Philip L. De Jager, Nilüfer Ertekin-Taner, David A. Bennett, Andrew J. Saykin, Annette Peters, Karsten Suhre, Rima Kaddurah-Daouk, Gabi Kastenmüller, Jan Krumsiek

## Abstract

Impaired glucose uptake in the brain is one of the earliest presymptomatic manifestations of Alzheimer’s disease (AD). The absence of symptoms for extended periods of time suggests that compensatory metabolic mechanisms can provide resilience. Here, we introduce the concept of a systemic ‘bioenergetic capacity’ as the innate ability to maintain energy homeostasis under pathological conditions, potentially serving as such a compensatory mechanism. We argue that fasting blood acylcarnitine profiles provide an approximate peripheral measure for this capacity that mirrors bioenergetic dysregulation in the brain. Using unsupervised subgroup identification, we show that fasting serum acylcarnitine profiles of participants from the AD Neuroimaging Initiative yields bioenergetically distinct subgroups with significant differences in AD biomarker profiles and cognitive function. To assess the potential clinical relevance of this finding, we examined factors that may offer diagnostic and therapeutic opportunities. First, we identified a genotype affecting the bioenergetic capacity which was linked to succinylcarnitine metabolism and significantly modulated the rate of future cognitive decline. Second, a potentially modifiable influence of beta-oxidation efficiency seemed to decelerate bioenergetic aging and disease progression. Our findings, which are supported by data from more than 9,000 individuals, suggest that interventions tailored to enhance energetic health and to slow bioenergetic aging could mitigate the risk of symptomatic AD, especially in individuals with specific mitochondrial genotypes.

## 1 Main

Dysregulation of bioenergetic pathways is a central feature of Alzheimer’s disease (AD), with detectable abnormalities occurring in the brain years prior to the symptomatic onset of AD dementia^1^. For example, studies investigating brain glucose uptake have consistently identified glucose hypometabolism as a presymptomatic metabolic manifestation of AD^2–4^. Additional evidence for early energetic dysregulation in AD comes from epidemiological studies, which have linked metabolic diseases, such as type 2 diabetes (T2D), and cardiovascular disease (CVD) to a significantly increased risk to develop AD later in life^5–9^. The delay between onset of symptomatic disease and apparent aberrations in energy pathways suggests the existence of a “bioenergetic capacity”, which can provide a temporary reserve that provides resilience from pathological symptoms of the disease.

Mitochondria are the essential cellular units of energy metabolism and are thus central to our proposed framework of energy-related resilience. They have been actively studied as potential targets for therapeutic intervention in AD^10,11^. Cellular energy supply through mitochondrial metabolism is fueled by three main routes that ultimately feed into the tricarboxylic acid (TCA) cycle: (1) Glucose catabolism to pyruvate, (2) beta-oxidation of fatty acids to acetyl-CoA, and (3) the degradation of proteins into glucogenic and ketogenic amino acids (**Fig. 1A**). These three routes are tightly controlled through a complex system of receptors and intracellular signaling that respond to the current metabolic state at both the organismal and tissue level^12^. In our study, we reduce the complexity of mitochondrial energetics by focusing on metabolic states under overnight fasting conditions. As a result, within the triad of energy pathways, dietary glucose metabolism assumes a marginal role (similar as in the above-mentioned state of reduced glucose uptake), while the metabolic routes of fatty acids and proteins becoming predominant.

**Fig. 1:**
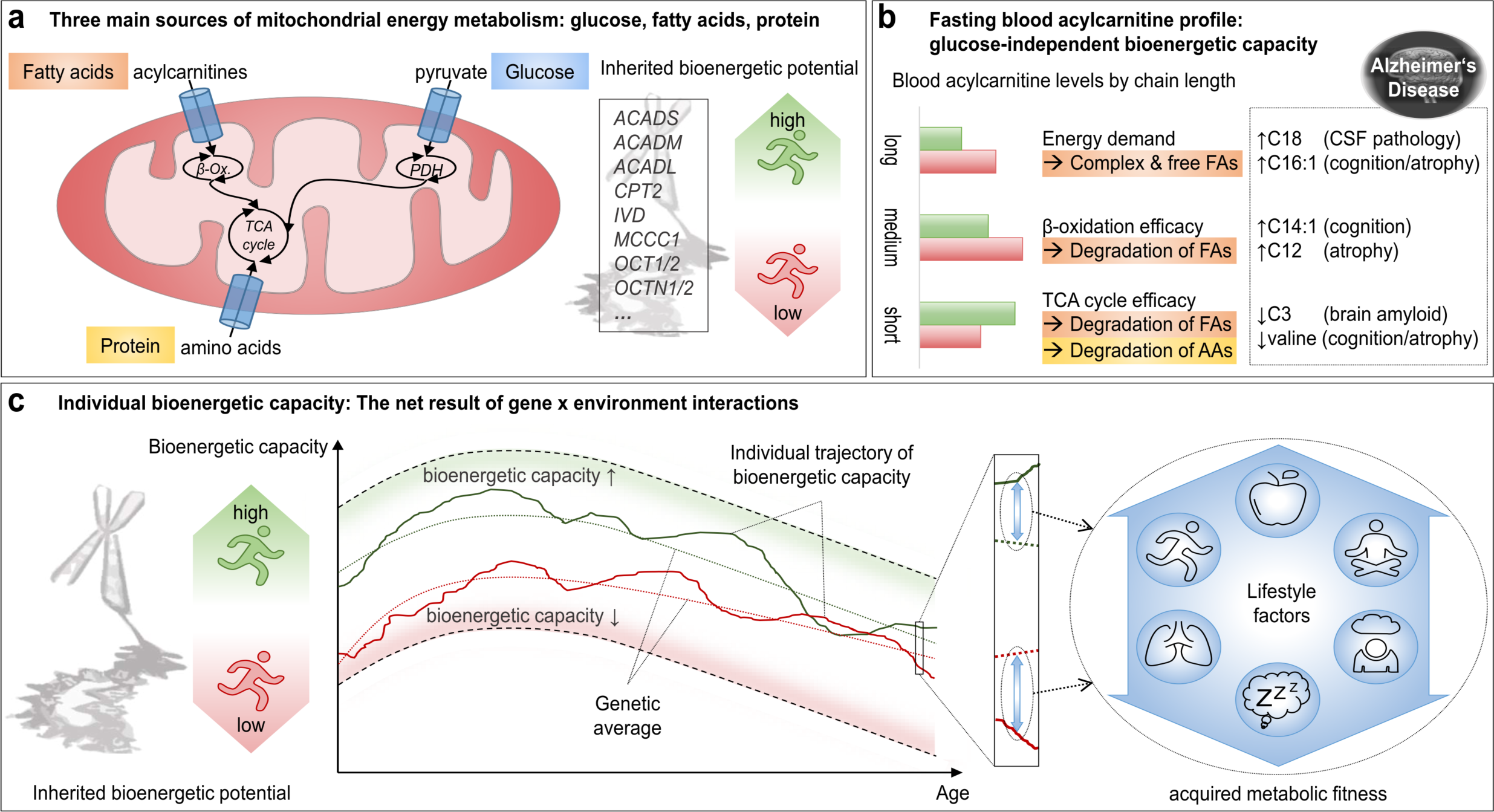
Concept of individual bioenergetic capacity mirroring impaired energy metabolism in the brain. **a.** The three main sources of mitochondrial energy metabolism: glucose, fatty acids, and proteins/amino acids, all of which ultimately feed into the TCA cycle. Common genetic variants in mitochondrial transporters and enzymes are assumed to define the inherited bioenergetic potential of each individual. Our study focuses on fasting individuals, largely removing the effect of dietary glucose and focusing on the fatty acid and protein routes. **b.** Chain length-specific role of acylcarnitines as readouts for the bioenergetic capacity through the functionality, activity and efficiency of mitochondrial energy metabolism; and examples of previously reported acylcarnitine level changes for AD-related phenotypes. **c.** Integrated concept of bioenergetic capacity as the age-specific result of inherited bioenergetic potential and acquired, modifiable metabolic functionality. Hypothetical trajectories for high and low inherited bioenergetic potential are shown, where deviations from the average are determined by modifiable lifestyle factors, such as physical activity, diet, health status, and other factors. Deviations from the overall population average are assumed to confer vulnerability or resilience to AD-related pathology and cognitive decline. Abbreviations: AD = Alzheimer’s disease; FAs: fatty acids; AAs: amino acids; CSF: cerebrospinal fluid; TCA: tricarboxylic acid; β-Ox.: beta-oxidation; PDH: Pyruvate dehydrogenase complex.

Acylcarnitines are a group of molecules whose blood levels provide accurate readouts specifically of the fatty acid and protein aspects of mitochondrial metabolism (**Fig. 1B**)^13^. They have long been used for the diagnosis of inborn errors of energy metabolism (IEEMs) through newborn screenings^13,14^. As a milder version of these deficiencies, genome-wide association studies (GWAS) have identified less penetrant single nucleotide polymorphisms (SNPs) that map to the same genes as in the IEEMs and show similar but weaker effects on blood acylcarnitine levels^15–18^. Acylcarnitine levels therefore serve as sensitive indicators of genetic influences on mitochondrial pathways. Beyond genetic variation, various acquired conditions influence mitochondrial pathways and are reflected in blood acylcarnitine levels. For example, increased levels of intermediate acylcarnitines from fatty acid and amino acid metabolism have been reported both in patients with T2D^19,20^ and in obese individuals with ketogenic branched-chain amino acid overload^21^. This suggests that the bioenergetic state by acylcarnitine levels might contain a modifiable component with consequences on disease risk. Interestingly, even during healthy aging, blood acylcarnitine levels reflect the age-dependent decrease in mitochondrial energetic capacity in both β-oxidation and TCA cycle pathways^22–24^. It has furthermore been proposed that blood acylcarnitine levels may be informative about the fatty acid oxidation status within the brain^25^.

Based on this combined evidence, we hypothesize that acylcarnitine profile in blood provide a proxy of an individual’s bioenergy capacity and thus their resilience buffer. We assume that deviations in this bioenergetic capacity from the normal population average significantly modulate the risk for disease outcomes, including neurodegenerative diseases like AD (**Fig. 1C**). In this study we show that acylcarnitine profiles can be used to: (1) Categorizing individuals by resilience status along their bioenergetic capacity. (2) Disentangle modifiable and genetic contributions to this resilience. (3) Integrate these profiles with genetic variation into a prognostic instrument that is predictive of future cognitive trajectories.

## 2 Results

### 2.1 Bioenergetic subgroups in AD and their determining factors

We performed subgroup Identification (SGI)^26^ on 1,531 ADNI participants using fasting serum profiles of 23 acylcarnitines as representative readouts of individual bioenergetic capacity. Study characteristics and acylcarnitine descriptions can be found in **Supplementary Tables 1 and 2**, respectively. Acylcarnitine levels were corrected for significant medication effects and ADNI study phase prior to clustering, but otherwise deliberately left uncorrected for any other potential confounders. SGI revealed a series of associations for cluster splits in the hierarchical tree relating to demographic variables, clinical diagnosis, and A-T-(N)-(C) measures^27^ (**Fig. 2a**).

**Fig. 2:**
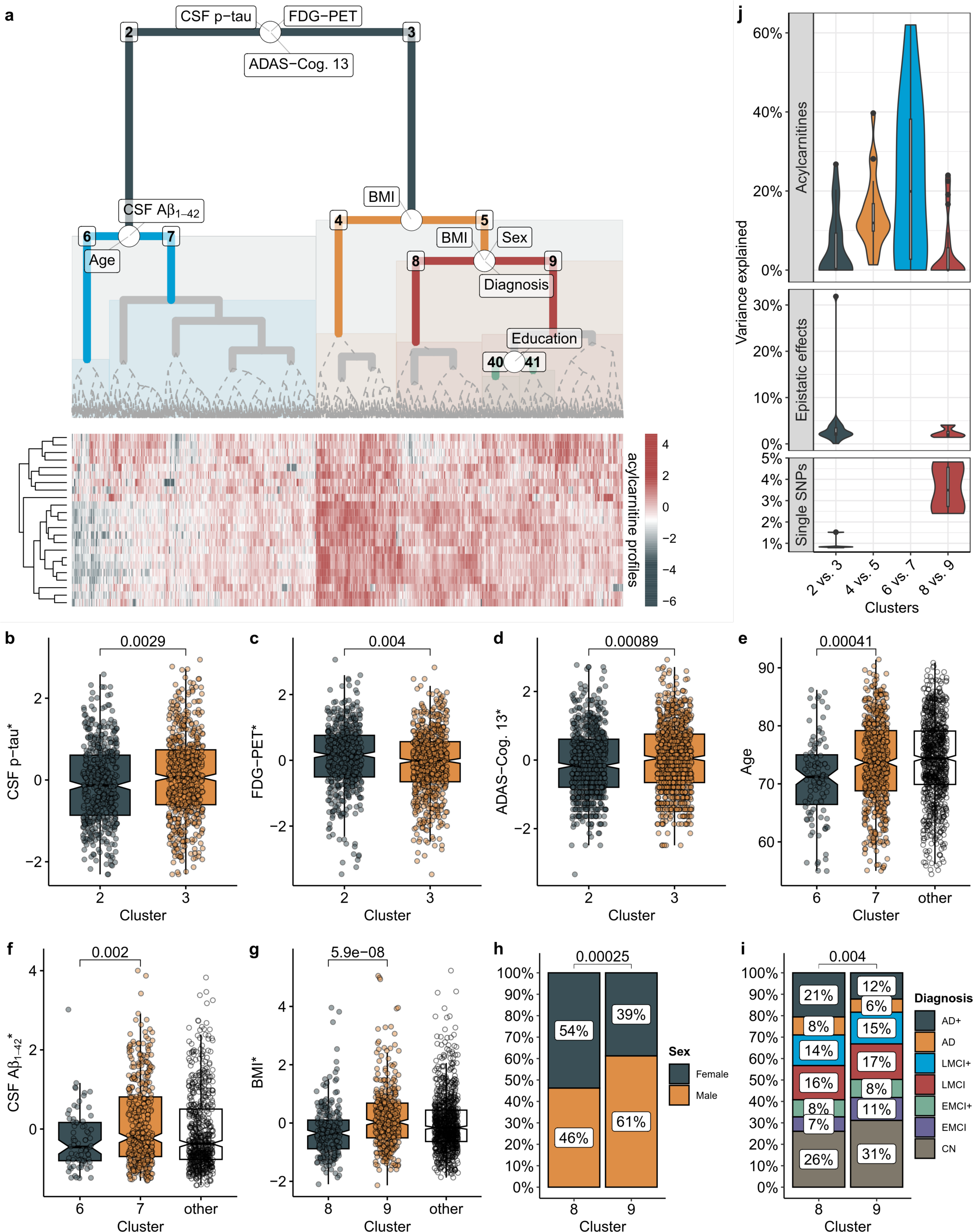
Acylcarnitine profiles stratify participants from the ADNI study in groups of different AD pathology. **a.** Acylcarnitine-based hierarchical clustering, with informative branches highlighted with solid-colored lines. Solid grey lines indicate cluster pairs that showed no significant associations. Clinical and demographic parameters at split points indicate significant differences between the individuals in the left and right subclusters (identified by number labels in the dendrogram) below that respective point. **b-d.** Individuals in cluster 2 have lower CSF p-tau levels, higher brain glucose uptake assessed by FDG-PET, and better cognitive function measured by the ADAS-Cog. 13 subscale compared to cluster 3. **e+f**. Further down on the left-hand side of the tree within favorable cluster 2, cluster 6 contains younger individuals with lower (worse) CSF Aβ_1-42_ compared to cluster 7. **g-i.** On the right hand-side of the tree, within cluster 3, cluster 8 contains a higher number of CSF amyloid-positive (indicated in the legend by ‘+’) individuals with clinical AD and a higher proportion of females compared to cluster 9. **j.** We investigated factors impacting subgroup division by examining both non-modifiable (acylcarnitine-related SNPs) and modifiable (adjusted acylcarnitine levels) factors. The results reveal a substantial amount (40-60%) of variance explained by genetics-corrected acylcarnitine levels, primarily medium- and long-chain, with overall rather minor contributions from genetic factors. The epistatic interaction between rs17806888 and rs924135, which explained ∼32% of the variance between clusters 2 and 3, is a notable exception. Both variants have been reported to significantly influence succinylcarnitine, highlighting a genetic link to the TCA cycle and amino acid-based energy metabolism. Variables marked with * have been centered to zero mean and scaled to unit variance. Abbreviations: CSF = cerebrospinal fluid, FDG-PET = Fluorodeoxyglucose Positron Emission Tomography, ADAS-Cog.13 = Alzheimer’s Disease Assessment Scale - Cognitive Subscale 13, BMI = body mass index.

The analysis split ADNI participants into two clusters, 2 and 3, at the top of the tree, with significantly different levels of p-tau in cerebrospinal fluid (CSF), glucose uptake measured by [^18^F] fluorodeoxyglucose-positron emission tomography (FDG-PET), and the 13-item AD assessment scale-cognitive subscale (ADAS-Cog. 13; **Fig. 2b-d, Supplementary Table 3**). Cluster 2 overall contained less affected individuals with significantly better cognitive function, higher brain glucose uptake, and lower CSF p-tau levels. This group was further divided into two distinct clusters: cluster 7, which on average was healthier with higher CSF Aβ_1-42_ levels and higher age, and cluster 6, a younger group showing signs of early pathological aging **(Fig. 2e+f)**. On the other side of the tree, within cluster 3 — which exhibited more progressed levels of Alzheimer’s disease biomarkers — there were further distinctions in clusters 8 and 9, which showed significant differences in the distribution of BMI, sex and diagnostic groups. (**Fig. 2g-i**). To rule out that these associations were driven by confounding factors, we repeated the analysis while adjusting for sex, age, body mass index (BMI), copies of APOE ε4, and years of education. All associations remained significant (**Supplementary Table 4**).

To investigate the factors potentially influencing subgroup division, we examined the variance explained by non-modifiable and modifiable factors for each branching point in the tree. Specifically, we examined acylcarnitine-related SNPs reported by Shin et al.^16^ as non-modifiable factors (SNP list is provided in Supplementary Table 5, replication in ADNI in Supplementary Table 6) and modifiable acylcarnitine levels (i.e., residuals adjusted for genetic and other non-modifiable factors) to better understand their impact on subgroup separation. Overall, sample clustering was mainly determined by the modifiable, covariate-adjusted acylcarnitine levels, which accounted for a significant portion of group differences with explained variance values ranging from 40% to 60% for different cluster branching points (**Fig. 2j**). Notably, the most substantial contributions were observed for medium- and long-chain acylcarnitines, highlighting a likely modifiable role for beta-oxidation function^28,29^ (**Supplementary Fig. 1**).

Genetic influences only accounted for a small proportion of variance (**Supplementary Table 7-9**), except for one epistatic interaction effect of two SNPs: The interplay of rs17806888 (mapped to *SUCLG2*) and rs924135 (mapped to *ABCC1*), both of which have been reported to affect succinylcarnitine levels^16^, explained 32% of the initial partitioning of the data into cluster 2 and cluster 3 at the top of the tree (**Fig. 2j** and **Supplementary Fig. 2**). When we examined relations to AD, we observed that both single acylcarnitine-related SNPs and epistatic models were not strongly linked to AD and its biomarkers, although some associations were independently confirmed in the MayoLOAD and ROS/MAP studies (**Supplementary Fig. 3** and **Supplementary Table 10**). The few genetic associations with AD parameters that we found were limited to influences on short-chain and dicarboxylic acylcarnitines. These findings indicate that the genetic link between AD, acylcarnitine pathways and the identified bioenergetic subgroups is primarily linked to the TCA cycle and amino acid-based energy metabolism.

In summary, these results suggest that individual bioenergetic capacity, represented by fasting serum acylcarnitine levels, can identify different groups of individuals. These include groups of study participants with less pronounced AD biomarker profiles, early pathological aging with decreased CSF Aβ_1-42_ levels and relatively low CSF p-tau levels, neurodegenerative processes accompanied by cognitive decline, and more advanced AD biomarker profiles. The stratification of study participants was primarily driven by the modifiable fraction of acylcarnitine levels involved in beta-oxidation, whereas the genetic component pointed towards the TCA cycle-related mechanisms.

### 2.2 Bioenergetic age correlates with Alzheimer’s disease pathology

Blood acylcarnitine levels have previously been described to significantly correlate with age^22^. Extending this concept, we hypothesized that incorporating acylcarnitine levels into a “bioenergetic age”, which might deviate from a person’s chronological age, can provide a single integrated readout of an individual’s bioenergetic capacity. After cross-normalizing the different cohort datasets for better comparability (**Methods** and **Supplementary Fig. 4**), we fitted a multivariable linear model that regresses age on the fasting serum levels of acylcarnitines in the KORA cohort (**Supplementary Fig. 5** and **Supplementary Table 11)**. KORA is a predominantly healthy, population-based cohort without prevalent AD cases, rendering it an appropriate reference for modeling the average aging process. The correlation between bioenergetic age and chronological age in KORA showed an r=62%. When applied to the ADNI cohort, correlation dropped to 28% (cognitively normal participants only), which was very similar to what we observed when replicating the bioenergetic age computation process in AGES-RS (r = 29%, **Supplementary Table 12**).

We then examined the different subgroups in ADNI concerning their bioenergetic age. First, we observed a gradual increase in bioenergetic age across the subgroups in the tree, ranging from cluster 6 with the lowest age, through clusters 7 and 5, to cluster 4 with the highest age (**Fig. 3a+b**). Furthermore, this analysis revealed a strong correlation between bioenergetic age and the first principal component of acylcarnitine levels, indicating that the primary axis of variation in the data and the clustering is associated with bioenergetic age (**Supplementary Fig. 6**). As we demonstrated in the previous section, the clustering seems primarily driven by modifiable factors, likely associated with individual beta-oxidation capacity. This finding thus suggests that bioenergetic age is also a modifiable factor, indicating that individuals could undergo interventions to transition into a different subgroup.

**Fig. 3:**
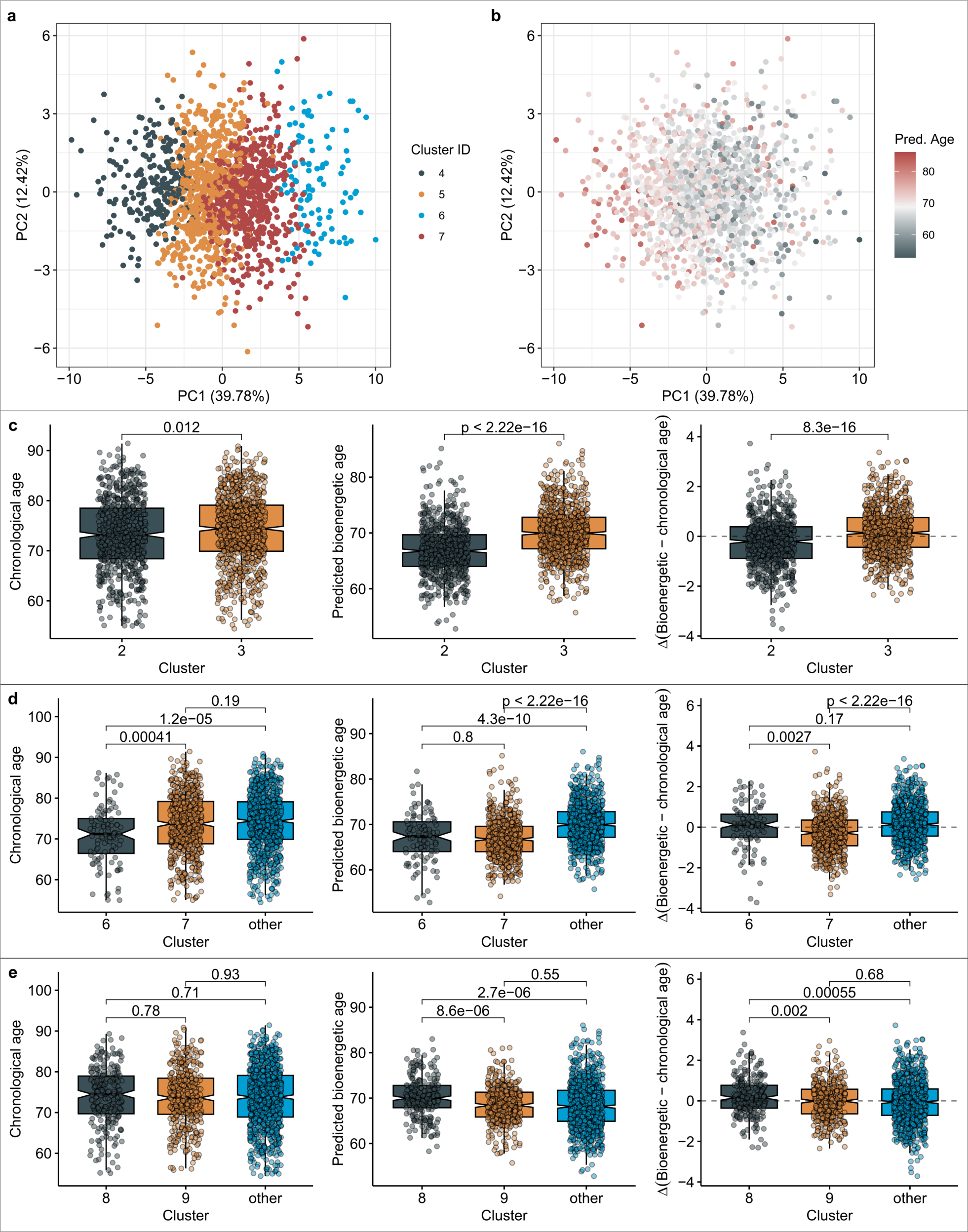
Bioenergetic age as a readout of bioenergetic capacity and determinant of bioenergetic subgroups. **a.** Principal component (PC) analysis of acylcarnitine profiles shows that the first PC expectedly follows the cluster structure from Fig. 2. **b.** Interestingly, the bioenergetic age predicted for the individuals within the dataset similarly corresponds with this cluster organization. **c.** Individuals in the pathologically healthier Cluster 2, although only slightly younger than those in Cluster 3 in terms of chronological age, display a significantly reduced bioenergetic age. **d**. Cluster 7, which is chronologically older than Cluster 6 but demonstrates favorable disease pathology, presents a bioenergetic age that is younger than their chronological age. This observation suggests that Cluster 7 may constitute a resilient subgroup of individuals. **e.** Similar to the previous two examples, individuals characterized by an advanced bioenergetic age exhibit more pronounced Alzheimer’s disease pathology compared to those with a younger bioenergetic age.

Bioenergetic age highlighted interesting relationships between subgroups and AD pathology throughout the tree (**Supplementary Table 13**). First, at the top split, we observed that while clusters 2 and 3 showed only a moderate difference in chronological age, there was a remarkable difference in bioenergetic age, with cluster 2 being substantially younger (**Fig. 3c**). This was in line with the observation that cluster 3 displayed progressed disease pathology compared to cluster 2. Further down the tree, bioenergetic age confirmed that the overall healthier cluster number 7 was substantially younger than the group’s chronological age, offering a potential explanation for their beneficial phenotypes (**Fig. 3d**). Importantly, this observed effect was not due to cluster 7 consisting of younger participants or a lower number of symptomatic individuals. Rather, we found that individuals within cluster 7 consistently exhibit the same chronological but significantly younger bioenergetic age than other participants irrespective of their diagnostic group (**Supplementary Fig. 7**). In clusters 8 and 9, we observe the same effect: Bioenergetically older individuals displayed increased AD pathology (**Fig. 3e**). This was further confirmed by significant associations of bioenergetic age with AD biomarkers across the A-T-(N)-(C) spectrum (**Supplementary Table 14**), and consistent findings for cognitive function and grey matter volume in AGES-RS (**Supplementary Table 15**).

In summary, we found that the acylcarnitine-based bioenergetic age metric, which we propose as a potential readout of a person’s bioenergetic capacity, showed strong associations with AD biomarkers beyond the natural aging process. Furthermore, our results suggest that bioenergetic age, which appears to be influenced by modifiable factors related to beta-oxidation function, can be a target for interventions to improve energy- and aging-related outcomes.

### 2.3 Bioenergetic age and succinylcarnitine-linked genotypes modulate future cognitive decline

We next assessed whether the bioenergetic age estimator can predict the trajectory of cognitive decline in AD, and whether this would allow us to identify subgroups of individuals that might particularly benefit from interventions.

We found that baseline bioenergetic age was significantly associated with the rate of cognitive decline in three different memory domains in the ADNI study (**Fig. 4a-c**). Specifically, bioenergetically younger individuals demonstrated a significantly slower decline over five years (**Supplementary Table 16**). This result was replicated using longitudinal diagnosis data from the AGES-RS study (**Fig. 4d, Supplementary Table 15**), providing further evidence that younger bioenergetic age is protective against cognitive decline.

**Fig. 4.**
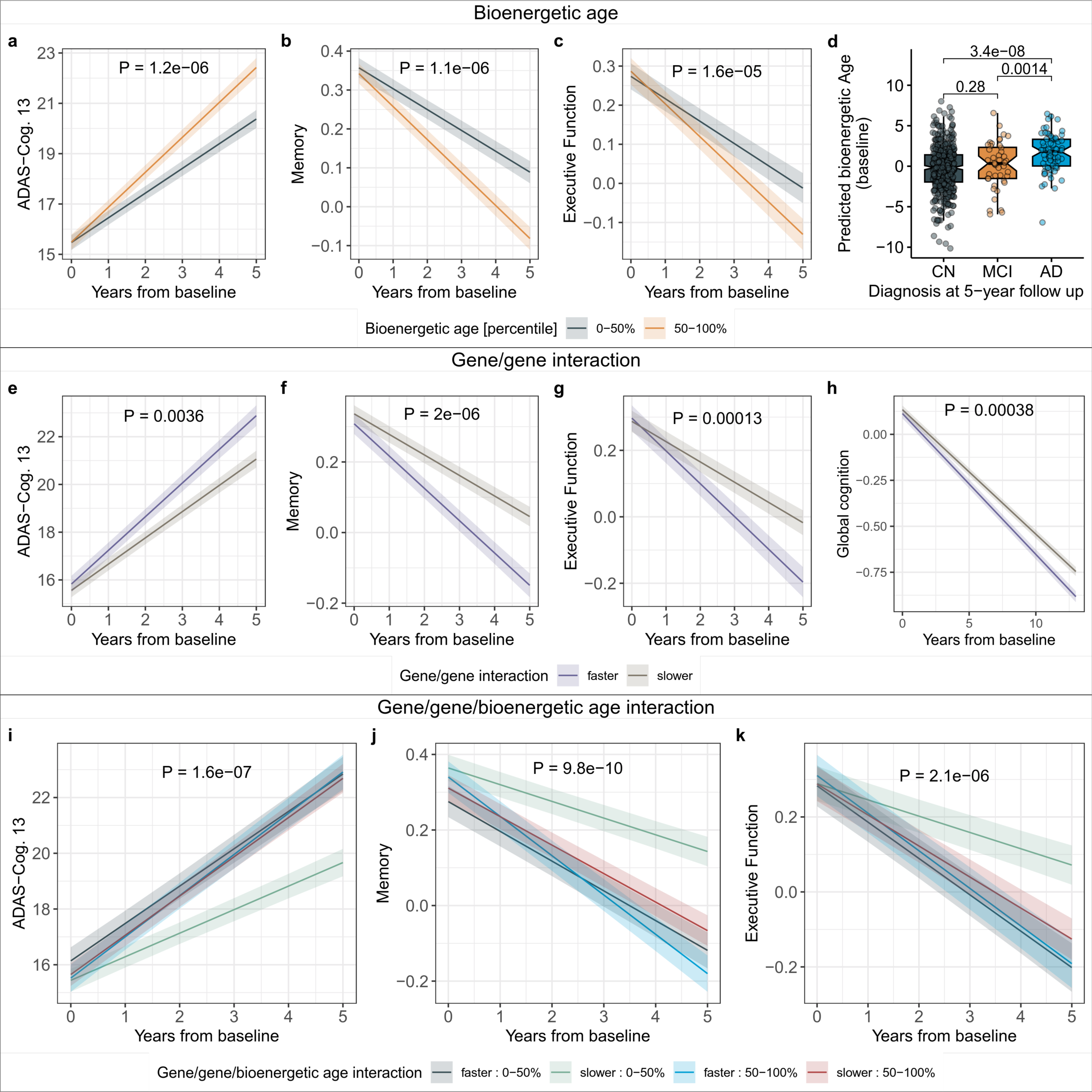
Bioenergetic age and succinylcarnitine-linked genotypes modulate the rate of cognitive decline. **a-c.** Bioenergetically younger individuals displayed a slower rate of cognitive decline compared to bioenergetically older individuals in the ADNI cohort. Median-split was only applied for visualization, reported P-values are for the continuous variable. **d.** Replication in the AGES study, by comparing bioenergetic age at baseline with clinical AD diagnosis after five years. **e-g.** Individuals with an unfavorable genotype configuration assessed by the combination of two SNPs, rs17806888 and rs924135, showed an accelerated rate of cognitive decline. **h.** Replication of the genetic signal in the ROS/MAP study. **i-k.** Interaction analysis: Only individuals with favorable bioenergetic age and favorable genotypes showed slower cognitive decline. This insinuates that individuals with unfavorable bioenergetic age but favorable genotypes could substantially benefit from targeted intervention.

We subsequently examined longitudinal cognitive trajectories in connection with the combination of SNPs rs17806888 and rs924135. These SNPs accounted for a substantial proportion of variance in the clustering (**Fig. 2j**). Since two SNPs can generate multiple genotype combinations, we identified a representative genotype by evaluating all possible groupings of allele combinations against the rest. We selected the grouping that had the most significant associations across three cognitive measures: ADAS-Cog 13, memory, and executive function (**Supplementary Table 17**). The genotype identified in this manner appears to significantly modulate cognitive decline (**Fig. 4e-g**). We attempted to replicate this grouping, which had similar genotype frequencies as in ADNI (59% vs. 58% in ROS/MAP; **Supplementary Fig. 8**), using trajectories of global cognition over 13 years in the ROS/MAP cohort. Interestingly, significant results only emerged when the analysis was restricted to individuals exhibiting clinically evident cognitive changes, such as a switch in cognitive classification from normal to MCI, or MCI to AD (**Fig. 4h**). These results suggest that specific genotype combinations related to short-chain acylcarnitines can influence the rate of future cognitive decline. Nonetheless, this effect might only be pertinent to individuals who are susceptible to cognitive alterations.

Finally, we examined the relationship between cognitive decline and a combination of bioenergetic age and the genotype groupings within the ADNI cohort. Remarkably, it seems that the protective influence of advantageous genotype combinations is limited to individuals with a younger bioenergetic age (**Fig. 4i-k**). This observation points to a highly interesting subgroup of individuals, those with beneficial genotypes but older bioenergetic age, who could potentially see considerable benefits from early interventions designed to decrease their predicted bioenergetic age. This subgroup constitutes approximately 30% of the ADNI participants. Replication analysis of this finding was not possible, since none of the available datasets except ADNI had combined fasting acylcarnitine measurements, genotyping, and longitudinal cognitive data.

## 3 Discussion

In this study, we investigated the association between Alzheimer’s disease (AD) pathology and bioenergetic capacity, approximated using fasting serum acylcarnitine levels. This bioenergetic capacity is influenced by a combination of modifiable and autosomal mitochondrial genes and naturally declines with age. Historically, acylcarnitine profiles have been widely used in clinical practice for detecting inborn errors of mitochondrial energy metabolism^30^. Moreover, numerous previous studies have shown the relevance of blood acylcarnitine levels and related mitochondrial pathways in metabolic disease and AD^31–37^. The present study combines these earlier findings with a translational framework of bioenergetic capacity with the potential for targeted interventions, based on collective evidence from more than 9,000 individuals.

Hierarchical clustering-based subgroup identification classified participants along the progression of their AD biomarkers. This stratification was predominantly driven by the modifiable fraction of acylcarnitine levels, specifically medium- and long-chain acylcarnitines associated with beta-oxidation function, accounting for 40-60% of variance explained. Notably, the division into the initial two main clusters was additionally and substantially influenced by an interaction between two genetic variants associated with succinylcarnitine, an intermediate of the TCA cycle and amino acid-based energy metabolism. These observations indicate that beta-oxidation might be a promising target for intervention, whereas targeting the TCA cycle may be complicated by complex genetic influences.

To integrate our findings into a unified score approximating an individual’s bioenergetic capacity, we derived a “bioenergetic age” metric, which showed a strong correlation with AD biomarkers, including brain glucose uptake, cognitive function, and disease progression. Furthermore, the two-SNP genotype related to succinylcarnitine, which influenced the clustering, was also predictive of cognitive decline. This is in line with previous findings reporting an association of genetic variation in the *SUCLG2* gene with cognitive decline^38^. Combined analysis of the bioenergetic age with the two-SNP model revealed that certain allele combinations appeared to result in resilience against cognitive decline, but only in individuals with a younger bioenergetic age.

Based on our results, we propose the following model of bioenergetic dysregulation in AD: As individuals age, their bioenergetic capacity decreases. This decrease accelerates with the onset of AD due to dysfunctional glucose uptake in the brain. The body then relies on alternative energy sources, primarily lipids via beta-oxidation and eventually protein-based energy production. Individuals with larger bioenergetic capacity, due to favorable genetics and maintained metabolic health, can temporarily compensate for these changes, resulting in resilience. However, when beta-oxidation can no longer provide sufficient energy, the body resorts to amino acid-based energy production. Only at that stage, amino acid metabolism will be negatively affected by genetic influences on related transporters and enzymes. This results in a ripple effect, increasing vulnerability to pathological processes and leading to accelerated disease progression. Overall, the bioenergetic capacity may thus be likened to a metabolic reserve providing a significant source of resilience against the disease.

Our key proposition – that the reduction of bioenergetic age will increase bioenergetic capacity and thus resilience in a genotype-specific manner – requires further validation in independent cohorts due to the rarity of studies that combine genetic, metabolomics, and longitudinal cognitive data. Once established as a robust marker of mitochondrial health, bioenergetic age combined with the two-SNP genotype could be utilized to select individuals for targeted interventions. Interventions to boost the bioenergetic capacity might include: (1) Low-carb or ketogenic diets, which directly influence mitochondrial beta-oxidation through nutritional lipids^39^, (2) physical activity, which is known to beneficially affect energy metabolism and mitochondrial fitness^40,41^, and (3) the use of drugs like Metformin, which was originally used to treat insulin resistance and type 2 diabetes, but recently has increasingly been shown to have additional beneficial effects, including the improvement of mitochondrial health^42,43^. Importantly, the bioenergetic age score could serve as a monitoring tool for such interventions, using established, cost-efficient, and fast acylcarnitine-measuring technologies, for example, based on dried blood spots^44^. While the ultimate long-term benefits of an intervention study can only be seen after decades, bioenergetic aging assessed through such minimally invasive acylcarnitine measurements is expected to predict success or failure of long-term interventions within a substantially shorter time frame.

## 4 Online Methods

### 4.1 ADNI study

The Alzheimer’s Disease Neuroimaging Initiative (ADNI) was launched in 2003 as a public-private partnership, led by Principal Investigator Michael W. Weiner, MD. The primary goal of ADNI has been to test whether serial magnetic resonance imaging (MRI), positron emission tomography (PET), other biological markers, and clinical and neuropsychological assessment can be combined to measure the progression of mild cognitive impairment (MCI) and early Alzheimer’s disease (AD). For up-to-date information, see www.adni-info.org. Written informed consent was obtained at enrollment, which included permission for analysis and data sharing. Consent forms were approved by each participating site’s institutional review board.

Data was obtained from the AMP-AD Knowledge Portal (https://adknowledgeportal.synapse.org), see **Data Availability Statement**, and the ADNI database at https://adni.loni.usc.edu. The AMP-AD Knowledge Portal is the distribution site for data, analysis results, analytical methodology, and research tools generated by the AMP-AD Target Discovery and Preclinical Validation Consortium and multiple Consortia and research programs supported by the National Institute on Aging.

Metabolomics data was available for 1,681 participants. Samples were profiled with the Biocrates p180 kit (Biocrates, Innsbruck, Austria). Metabolomics data processing largely followed a previously published protocol^35^: Of the 186 metabolites covered by the platform, four were removed due to technical issues, leaving a total of 182 metabolites for further analysis. Samples were distributed across 23 plates. Each plate included NIST Standard Reference samples. 22 metabolites with large numbers of missing values (> 40%) were excluded and plate batch effects were removed by cross-plate mean normalization using NIST sample metabolite concentrations. The sample set moreover contained blinded duplicated measurements for 19 samples (ADNI-1) and blinded triplicated measurements for 17 samples (ADNI-GO and −2) distributed across plates. These duplicated and triplicated study samples were used to remove 20 metabolites with coefficients of variation >20% or intra-class correlation <65%. Biological replicates were then averaged, and non-fasting participants (n=108) were excluded. We imputed missing metabolite data using half the value of the lower limit of detection per metabolite, log2-transformed metabolite concentrations, centered and scaled distributions to a mean of zero and unit variance and winsorized single outlying values to 3 standard deviations. Mahalanobis distance was used for the detection of multivariable subject outliers. Applying a critical Chi-square value of P < 0.01 resulted in the removal of 42 samples. Finally, metabolites were adjusted for significant medication effects using stepwise backwards selection (for details see Toledo et al.^33^). The final dataset contained 140 metabolite measurements, covering 23 acylcarnitine species (**Supplementary Table 2**), for 1,531 individuals.

Representative phenotypes across A-T-(N)-(C) measures^27^ were used as clinical phenotypes, including baseline levels of CSF amyloid β_1-42_ (CSF Aβ_1-42_, **A**), CSF p-tau (**T**), ROI-based FDG-PET measures of average glucose uptake across the left and right angular, left and right temporal, and bilateral posterior cingulate regions (**N**), and ADAS-Cog. 13 scores (**C**). Diagnostic groups were coded as follows: 1 = cognitively normal (CN) individuals and individuals with subjective memory complaints (SMC); 2 = early MCI (EMCI); 3 = EMCI with CSF Aβ_1-42_ pathology; 4 = late MCI (LMCI); 5 = LMCI with CSF Aβ_1-42_ pathology; 6 = AD cases; 7 = AD cases with CSF Aβ_1-42_ pathology. Sex, age, BMI, years of education and copies of *APOE* ε4 were included as covariates in cross-sectional association tests. Epistatic analyses were additionally adjusted for ADNI study phase. Longitudinal analyses of cognitive trajectories were adjusted for age and diagnosis at baseline, sex, copies of *APOE* ε4, education, and ADNI study phase. Longitudinal analyses were restricted to five years of follow up to retain statistical power (minimal n = 378).

Whole genome-genotyping was available for 1,548 ADNI participants, with 1,378 participants having overlapping metabolomics data. Genotyping data were collected using the Illumina Human 610-Quad, HumanOmni Express, and HumanOmni 2.5M BeadChips. Pre-imputation quality control procedures included filtering for SNP call rate < 95%, Hardy-Weinberg equilibrium test p-value < 1 x 10^-6^, minor allele frequency < 1%, participant call rate < 95%, and discordance between reported and inferred sex. Non-Hispanic Caucasian participants were selected using HapMap 3 genotype data and multidimensional scaling analysis. Genotype imputation was performed for each BeadChip type separately using the Haplotype Reference Consortium (HRC) reference Panel r1.1.

### 4.2 ROS/MAP study

The Religious Order Study (ROS) and the Rush Memory and Aging Project (MAP) studies^45^ are longitudinal cohort studies of aging and AD, conducted by the Rush Alzheimer’s Disease Center and designed to be used in joint analyses to maximize sample size. Both studies were approved by an Institutional Review Board at Rush University Medical Center. All participants signed an informed consent and a repository consent to allow their biospecimens and data to be used for ancillary studies. Further, all participants signed an Anatomic Gift Act for organ donation for research. More details can be found at www.radc.rush.edu.

Imputed genome-wide genotype data for 2,059 study participants was obtained from the AMP-AD Knowledge Portal (https://adknowledgeportal.synapse.org), see **Data Availability Statement**. A description of this data, including quality control procedures and imputation, was previously published^46^. We further included phenotypic data on clinical diagnosis at death, global cognition during lifetime, amyloid-β and paired helical filament (PHF)-tau protein load in brain tissue, global burden of AD neuropathology (mean of neuritic plaques, diffuse plaques, and neurofibrillary tangles).

Epistatic analyses were adjusted for sex, age at death, education, post-mortem interval, and number of copies of *APOE* ε4 as covariates. Longitudinal analysis of cognitive trajectories was adjusted for baseline age (instead of age at death) and clinical diagnosis, while post-mortem interval was omitted here. Longitudinal analysis was restricted to thirteen years of follow up, as at this time point we had a similar sample size left (n = 368) as in the longitudinal analyses in ADNI.

### 4.3 MayoLOAD study

The Mayo study of late-onset AD (MayoLOAD)^47^ is a case/control study from three different series: Mayo Clinic Jacksonville, Mayo Clinic Rochester and Mayo Clinic Brain Bank series. The study was approved by the appropriate institutional review board and appropriate informed consent was obtained from all participants. Preprocessed genotype data for 2,067 participants were obtained from the AMP-AD Knowledge Portal (https://adknowledgeportal.synapse.org), see **Data Availability Statement**. Briefly, samples with call rates <90% were removed. In addition, samples were discarded based on mismatch between inferred and reported sex. Further, samples were filtered based on inferred relatedness to ensure that the resultant sample set represents unrelated individuals. SNPs with call rates <90%, minor allele frequencies <0.01, and/or Hardy-Weinberg P values <0.001 were eliminated. Total genotyping rate after filtering was 99.2%. Genotypes were then imputed using the 1000 genomes phase 3 reference panel^48^ by first prephasing genotypes using SHAPEIT2 (v 2.12)^49^ and then imputing using IMPUTE2 (v2.3.2)^50^.

Data on clinical diagnosis was numerically coded into four categories: 1 = controls with no evidence for AD-related neuropathology; 2 = clinically normal controls without neuropathology assessment; 3 = Clinical AD without neuropathology confirmation; 4 = clinical diagnosis of AD dementia with neuropathology-confirmed AD. Covariates for the study included sex, age at death, and number of copies of *APOE* ε4.

### 4.4 KORA study

The Cooperative Research in the Region of Augsburg (KORA) study is a population-based sample from the general population living in the region of Augsburg, Southern Germany^51^. Here we used data from the KORA F4 study, the first follow-up examination of KORA S4 in 2006-2008. The study, including the protocols for subject recruitment and assessment and the informed consent for participants, was reviewed and approved by the local ethical committee (Bayerische Landesärztekammer).

Metabolomics data was available for 3,029 predominantly healthy participants with data on fasting serum acylcarnitine levels, sex, age, and BMI. Metabolic profiling was conducted using the Biocrates p150 kit (Biocrates, Innsbruck, Austria), a precursor of the p180 kit where measurements of acylcarnitines are performed analogously as on the p180 kit. A detailed description of processing of metabolomics data is provided in Mittelstrass et al.^52^. Briefly, similar to the ADNI procedure, filters for coefficient of variation (< 25%), missingness (< 10%), and correlation of repeated measurements (> 50%) were applied to remove metabolites of limited measurement quality. Multivariable subject outliers were identified using the Mahalanobis distance, remaining missing values were imputed, and data was log-transformed for subsequent analyses. Data for 22 out of 23 acylcarnitines investigated in ADNI (**Supplementary Table 2**) were available here, with the exception being levels of C4:1, which was not reliably measured in KORA.

### 4.5 AGES-Reykjavik study

The Age, Gene/Environment Susceptibility-Reykjavik Study (AGES-RS) is an epidemiologic study focusing on four biologic systems: vascular, neurocognitive (including sensory), musculoskeletal, and body composition/metabolism^53^. AGES-RS was approved by the National Bioethics Committee in Iceland that acts as the Institutional Review Board for the Icelandic Heart Association (approval number: VSN-00-063), and by the National Institute on Aging Intramural Institutional Review Board. A multistage consent is obtained in AGES-RS to cover participation, use of specimens and DNA, and access to administrative records.

Fasting serum-based measurements for the 23 acylcarnitines generated with the Biocrates p180 kit (Biocrates, Innsbruck, Austria) were available for 575 AGES-RS participants at baseline, of which 544 had a follow-up diagnostic assessment after 5 years. As in ADNI, data was batch-normalized, log2-transformed, centered and scaled, and adjusted for medication effects. All analyses in AGES-RS were adjusted for age, sex, education, and copies of *APOE* ε4. In the analysis of grey matter volume, intracranial volume was included as additional covariate.

### 4.6 Subgroup identification analysis (SGI)

We used the SGI software package^26^ for automatic subgroup identification. For the present study, the analysis consisted of the following three steps. First, the data matrix was standardized (mean 0, standard deviation 1 for each metabolite) before analysis. The samples were then hierarchically clustered based on their acylcarnitine profiles using the Euclidean distance metric and Ward linkage^54^. This resulted in a dendrogram, i.e., a binary tree that provides the hierarchical structure of sample similarities. Second, we performed association analysis with AD-related phenotypes and the 32 acylcarnitine-associated SNPs. Each branching point (BP) in the dendrogram provides two subgroups of participants, from the top BP that separates the entire dataset into two parts over smaller clusters down to the bottom where subgroups consist of only a few samples each. For each BP, an association test of the respective left vs. right cluster was performed using linear regression for A-T-(N)-(C) measures, and ordinal regression for copies of *APOE* ε4 and the numeric coding of diagnosis. To avoid low-powered statistical tests, BPs were only tested if both of the two underlying subgroups were larger than N = 77 samples (valid cluster pairs), which corresponds to 5% of all samples. Third, we adjusted p-values for multiple testing by correcting by the number of valid cluster pairs *s*=11. Since a dendrogram’s clusters are nested, non-overlapping groups, the *s* statistical tests performed are strictly independent. Thus, control of the family-wise error rate can be achieved using a Bonferroni-like multiple testing correction by adjusting each p-value by a factor of *s* (corresponding to an adjusted threshold of p ≤ 4.55 x 10^-3^).

### 4.7 Collection of SNPs and SNP combinations associated with blood acylcarnitine levels

We obtained a list of 32 acylcarnitine-associated SNPs (**Supplementary Table 5**) from a large GWAS study performed on a total of 7,824 individuals from the KORA and TwinsUK studies^16^. Generalizability of effects of these SNPs to ADNI was tested using a targeted genetic association screening, where we tested for influences of all 32 SNPs against all 23 acylcarnitine species assuming an additive genetic model. Only age and sex were included as covariates, following the protocol of the reference study. Associations were considered to be significant if they had an FDR-adjusted p-value ≤ 0.05.

The reference GWAS further lists multi-SNP combinations of one lead SNP per associated locus that in concert explained the largest fraction of the heritable population variance of single acylcarnitine concentrations in blood. We used these multi-SNP combinations in epistatic modeling.

### 4.8 Epistatic analysis

To determine multi-locus epistatic associations with the clustering and AD-related outcomes, we ran an epistasis model using SNP combinations for each acylcarnitine with significant associations in more than one locus. For these multi-locus genetic models, we combined SNP genotypes in all combinations into aggregated genotypes *h* to test pure genetic interactions; for example, if SNP1 has alleles TA for an individual, and SNP2 has alleles CC, then the aggregated genotype is TACC. Since each SNP can have up to three different genotypes (major allele homozygote, heterozygote, minor allele homozygote), the aggregation of N SNPs can yield up to 3^N^ combinations. For each outcome (cluster pairs and AD-related phenotypes) *y* and aggregated genotype *h*, we then computed the following two statistical models: the base model *M_1_* regressing the outcome in question (dependent variable) on the set of confounders only; and the full model *M_2_* that includes *h* in addition to confounders. The models use ordinal regression with log-log link functions and logistic regression for binary outcomes. The genetic variable *h* is treated as a factor and is thus expanded into a binary indicator variable for each factor level during model fitting. If a factor level is observed in less than 10 participants, it was omitted due to lack of statistical power. To control for spurious effects of overly fractionated factor variables with many different SNP combinations, we additionally capped the maximum number of degrees of freedom (DF). This was achieved by imposing a ridge-type penalty following the suggestions described by Harrell^55^ and setting the maximum number of DFs to 15. Statistical significance was finally assessed using a likelihood ratio test between *M_1_* and *M_2_*.

### 4.9 Analysis of explained variance

Variance between cluster pairs explained by significant genetic effects (both for single SNPs and multi-SNP combinations) was estimated using McKelvey’s measure, which has been described as a robust approach for logistic regression models^56^. To estimate the variance explained by each of the 23 acylcarnitines, we calculated a linear mixed model including a random intercept for each branching point, as implemented in the variancePartition R package^57^. To account for potential confounding through non-modifiable factors, we included all significant SNPs, the interaction between rs17806888 and rs924135, as well as age, sex, BMI, and Education as covariates in the model. Analysis of explained variance was restricted to cluster pairs for which we observed significant differences in AD-related phenotypes or demographic variables.

### 4.10 Computation and analysis of bioenergetic age

For learning the linear predictor of chronological age in the KORA F4 study, we extracted all acylcarnitine measurements (n = 22) that were also available in ADNI and AGES-RS. We then extracted a reference set of healthy female individuals in both KORA, ADNI, and AGES-RS. Selection parameters in KORA besides female sex included age 60-72 years and BMI 23.31-29.72 kg/m². In ADNI and AGES-RS we included all cognitively normal females with an age of 72 years or younger. Acylcarnitines were then centered and scaled in each reference set separately and filtered for multivariate outliers (n = 3; all ADNI) based on the Mahalanobis distance (see more details in the description of ADNI data). The remaining reference subjects were used to rescale z-scored acylcarnitine concentrations in the respective complete cohorts. After normalizing acylcarnitine levels to these reference groups, we calculated a linear regression model using the reference-transformed 22 acylcarnitines to predict chronological age in KORA. To investigate robustness of the model, we performed 10-fold, 3 times repeated cross-validation (**Supplementary Table 11**). We then applied this model to ADNI and AGES-RS using reference-transformed acylcarnitine levels. In this analysis, we did not adjust acylcarnitine levels for ADNI study phase, as participants in ADNI-GO/2 were on average 2.65 years younger than participants in ADNI-1 (P = 5.76 x 10^-13^), such that adjustment would have confounded the reference set-based rescaling. The z-scored difference of chronological age and bioenergetic age was derived by subtracting their z-scored transformations, centering to zero and scaling to unit variance. Differences between ADNI cluster pairs associated with AD-related phenotypes and the three age measures (chronological age, predicted bioenergetic age, and their delta) were assessed using linear regression without adjustment for any additional variables. Cross-sectional associations of predicted bioenergetic age with A-T-(N)-(C) measures in ADNI, as well as with total grey matter volume, cognition and clinical diagnosis in AGES-RS were tested using linear regression while adjusting for all relevant covariates (see study-specific sections), including chronological age.

### 4.11 Analysis of longitudinal cognitive trajectories

For analyses of cognitive trajectories in ADNI, we included the composite ADNI scores for memory (ADNI-MEM)^58^ and executive function (ADNI-EF)^59^ in addition to the ADAS-Cog. 13, which have been described to be more sensitive to subtle cognitive changes and have been used in studying resilience to AD before^60^. We tested longitudinal associations with predicted bioenergetic age using linear mixed effects models with cognitive scores as dependent variables. The explanatory variable of interest was the interaction of bioenergetic age and time. Models were adjusted for relevant covariates (see description of ADNI data), including chronological age at baseline, and allowed for random intercepts for each participant. For binarization of the SNP combination of rs17806888 and rs924135 into two groups (slower vs. faster progression of cognitive decline), we used the same linear mixed effect model (replacing bioenergetic age with genotype aggregates) and iterated over all binary combinations of aggregated genotypes. We removed aggregates with less than 20 observations to avoid spurious associations, leaving us with six aggregated genotypes. We then selected the combination that showed the highest significance across the three cognitive scores using Fisher’s sum of logs method^61^. The three-way interaction analysis was performed in the same way (explanatory variable of interest: time x bioenergetic age x SNP grouping), while for the calculation of the association p-value, we used an ANOVA, where the reduced model omitted the interaction with time. In AGES-RS, we used standard linear regression to test for an association between baseline predicted bioenergetic age and pheno-conversion after five years, while adjusting for relevant covariates. To replicate the genotype grouping obtained in ADNI in ROS/MAP, we applied the same (in terms of predictors/covariates) linear mixed effects model as in ADNI using global cognition as outcome. As replication in the full subset of the ROS/MAP cohort available here failed, we selected a subset of n = 1,081 participants (total 1,936 with cognitive and genetic data available) where a clinically relevant change in cognitive status (from cognitive normal to MCI or AD, or from MCI to AD) was noticed in any follow-up visit (1-26 years, mean time until change = 5.71 years).

## Data availability statement

**KORA** data can be accessed upon request at https://helmholtz-muenchen.managed-otrs.com/external/.

**AGES-RS** data can be accessed upon request according to informed consent at https://hjarta.is/en/research/ages-phase-1/.

All other omics datasets are available via the AD Knowledge Portal (https://adknowledgeportal.org). The AD Knowledge Portal is a platform for accessing data, analyses, and tools generated by the Accelerating Medicines Partnership (AMP-AD) Target Discovery Program and other National Institute on Aging (NIA)-supported programs to enable open-science practices and accelerate translational learning. The data, analyses and tools are shared early in the research cycle without a publication embargo on secondary use. Data is available for general research use according to the following requirements for data access and data attribution (https://adknowledgeportal.org/DataAccess/Instructions).

For access to content described in this manuscript see:

**ADNI** metabolomics data from the AbsoluteIDQ-p180 kit is available at the AD Knowledge Portal under https://doi.org/10.7303/syn5592519 (ADNI-1) and https://doi.org/10.7303/syn9705278 (ADNI-GO/-2), the full complement of clinical and demographic data for the ADNI cohorts are hosted on the LONI data sharing platform and can be requested at http://adni.loni.usc.edu/data-samples/access-data/.

**ROS/MAP** imputed genotype data is available at the AD Knowledge Portal under https://doi.org/10.7303/syn3157329, study meta-data, basic covariates and clinical variables are available under https://doi.org/10.7303/syn3157322. The full complement of clinical and demographic data for the ROS/MAP cohorts are hosted on the RADC data sharing platform and can be requested at www.radc.rush.edu.

**MayoLOAD** genotyping data is available at the AD Knowledge Portal under https://doi.org/10.7303/syn3157238, study meta-data and covariates are available under https://doi.org/10.7303/syn3205821.6.

## Acknowledgements

This work was supported by the National Institutes of Health/the National Institute of Aging through grants RF1AG058942, RF1AG059093, U01AG061359, U19AG063744 and R01AG069901. In addition, the following funding sources are acknowledged:

**ADNI:** Data collection and sharing for this project was funded by the Alzheimer’s Disease Neuroimaging Initiative (ADNI) (National Institutes of Health Grant U01 AG024904) and DOD ADNI (Department of Defense award number W81XWH-12-2-0012). ADNI is funded by the National Institute on Aging, the National Institute of Biomedical Imaging and Bioengineering, and through generous contributions from the following: AbbVie, Alzheimer’s Association; Alzheimer’s Drug Discovery Foundation; Araclon Biotech; BioClinica, Inc.; Biogen; Bristol-Myers Squibb Company; CereSpir, Inc.; Cogstate; Eisai Inc.; Elan Pharmaceuticals, Inc.; Eli Lilly and Company; EuroImmun; F. Hoffmann-La Roche Ltd and its affiliated company Genentech, Inc.; Fujirebio; GE Healthcare; IXICO Ltd.; Janssen Alzheimer Immunotherapy Research & Development, LLC.; Johnson & Johnson Pharmaceutical Research & Development LLC.; Lumosity; Lundbeck; Merck & Co., Inc.; Meso Scale Diagnostics, LLC.; NeuroRx Research; Neurotrack Technologies; Novartis Pharmaceuticals Corporation; Pfizer Inc.; Piramal Imaging; Servier; Takeda Pharmaceutical Company; and Transition Therapeutics. The Canadian Institutes of Health Research is providing funds to support ADNI clinical sites in Canada. Private sector contributions are facilitated by the Foundation for the National Institutes of Health (www.fnih.org). The grantee organization is the Northern California Institute for Research and Education, and the study is coordinated by the Alzheimer’s Therapeutic Research Institute at the University of Southern California. ADNI data are disseminated by the Laboratory for Neuro Imaging at the University of Southern California.

Data collection and sharing for this project was further funded by the Alzheimer’s Disease Metabolomics Consortium (National Institute on Aging R01AG046171, RF1AG051550 and 3U01AG024904-09S4).

**ROS/MAP:** Study data were provided by the Rush Alzheimer’s Disease Center, Rush University Medical Center, Chicago. Data collection was supported through funding by NIA grants P30AG10161 (ROS), R01AG15819 (ROSMAP; genomics and RNAseq), R01AG17917 (MAP), R01AG30146, R01AG36042 (5hC methylation, ATACseq), RC2AG036547 (H3K9Ac), R01AG36836 (RNAseq), R01AG48015 (monocyte RNAseq) RF1AG57473 (single nucleus RNAseq), U01AG32984 (genomic and whole exome sequencing), U01AG46152 (ROSMAP AMP-AD, targeted proteomics), U01AG46161(TMT proteomics), U01AG61356 (whole genome sequencing, targeted proteomics, ROSMAP AMP-AD), the Illinois Department of Public Health (ROSMAP), and the Translational Genomics Research Institute (genomic). Additional phenotypic data can be requested at www.radc.rush.edu.

**MayoLOAD:** Study data were provided by the Mayo Clinic Alzheimer’s Disease Genetic Studies, led by Dr. Nilüfer Ertekin-Taner and Dr. Steven G. Younkin, Mayo Clinic, Jacksonville, FL using samples from the Mayo Clinic Study of Aging, the Mayo Clinic Alzheimer’s Disease Research Center, and the Mayo Clinic Brain Bank. Data collection was supported through funding by NIA grants P50 AG016574, R01 AG032990, U01 AG046139, R01 AG018023, U01 AG006576, U01 AG006786, R01 AG025711, R01 AG017216, R01 AG003949, NINDS grant R01 NS080820, CurePSP Foundation, and support from Mayo Foundation.

**KORA:** The KORA study was initiated and financed by the Helmholtz Zentrum München – German Research Center for Environmental Health, which is funded by the German Federal Ministry of Education and Research (BMBF) and by the State of Bavaria.

**AGES-RS:** The AGES-RS study has been funded by NIH contract N01-AG-12100, the NIA Intramural Research Program, Hjartavernd (the Icelandic Heart Association) contract HHSN271201200022C, and the Althingi (the Icelandic Parliament).

Funding sources had no role in study design, the collection, analysis and interpretation of data, the writing of the report, or in the decision to submit the article for publication.

## Author contributions

Conceptualization and methodology, M.A., M.B., G.K. and J.K.; data analysis, M.A., M.B., and T.W.; metabolomics data generation and QC, M.A., G.K., R.W.S., J.A., A.P., V.G., L.J.L., K.S. and R.K.D; genotype data generation and QC, M.A., P.L.D.J., N.E.T., D.A.B., K.N., and A.J.S.; visualization, M.A., M.B., G.K., and J.K.; supervision, R.K.D., A.J.S., M.A., G.K., J.K., P.L.D.J. N.E.T., D.A.B., V.G., L.J.L., K.S. and P.M.D.; funding acquisition, R.K.D., A.J.S., M.A., G.K., P.L.D.J., N.E.T., D.A.B., and P.M.D.; writing—original draft, M.A., M.B., G.K., J.K., and P.M.D.; results interpretation, M.A., M.B., K.S., G.K., and J.K.; writing—review and editing, all authors. All authors read and approved the final manuscript.

