## Supplementary Figures for "Individual bioenergetic capacity as a potential source of resilience to Alzheimer’s disease"

### Supplementary Figure 1

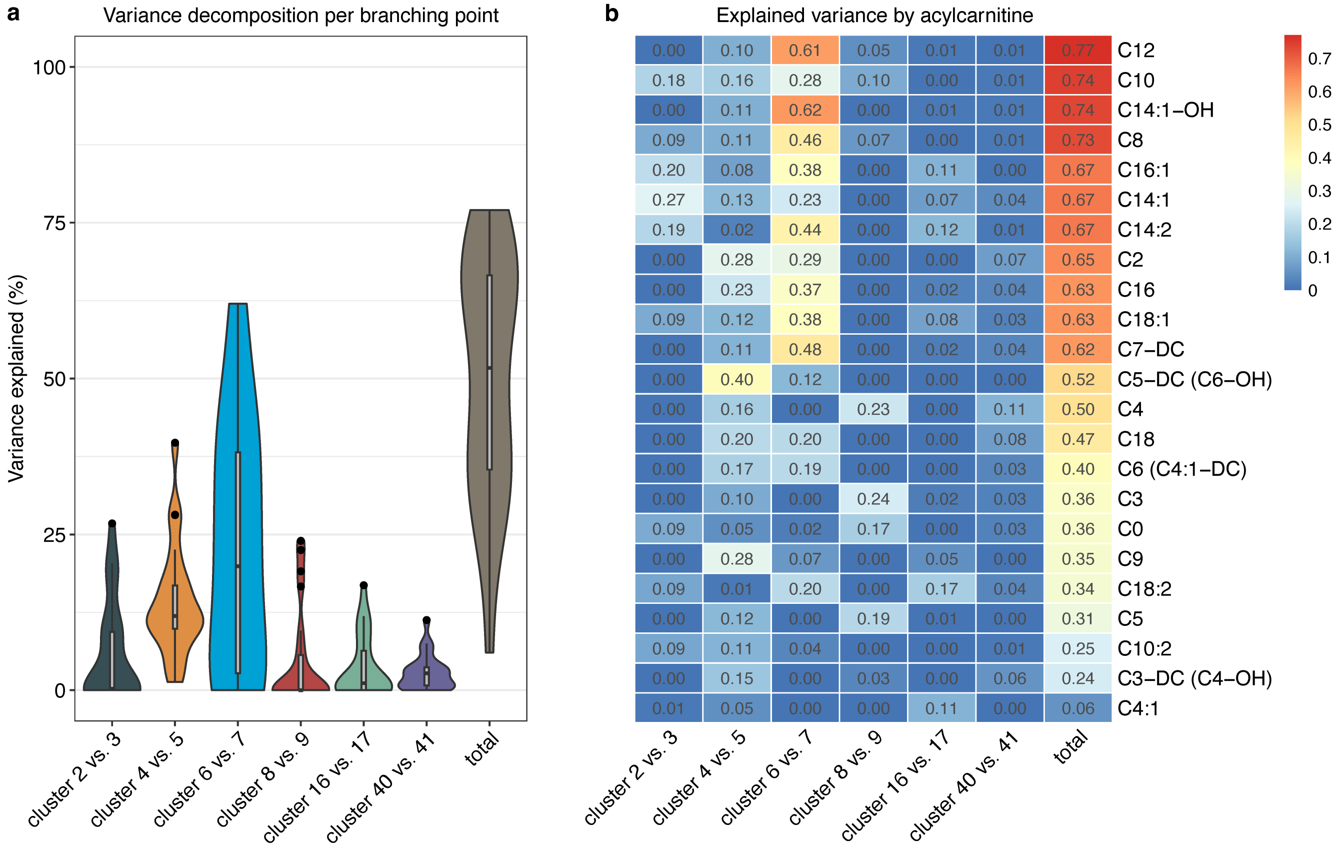

**Variance of branching points in the clustering explained by modifiable fractions of acylcarnitine levels.** This figure expands on the data provided in Figure 2j, illustrating estimates of explained variance for the clustering of acylcarnitines across all cluster pairs that showed associations to Alzheimer's Disease (AD) or demographics. Two distinct visualizations are presented: (**a**) Violin plots showcasing distributions across acylcarnitines; and (**b**) heatmap with individual metabolite-level estimates.

#### Supplementary Figure 2

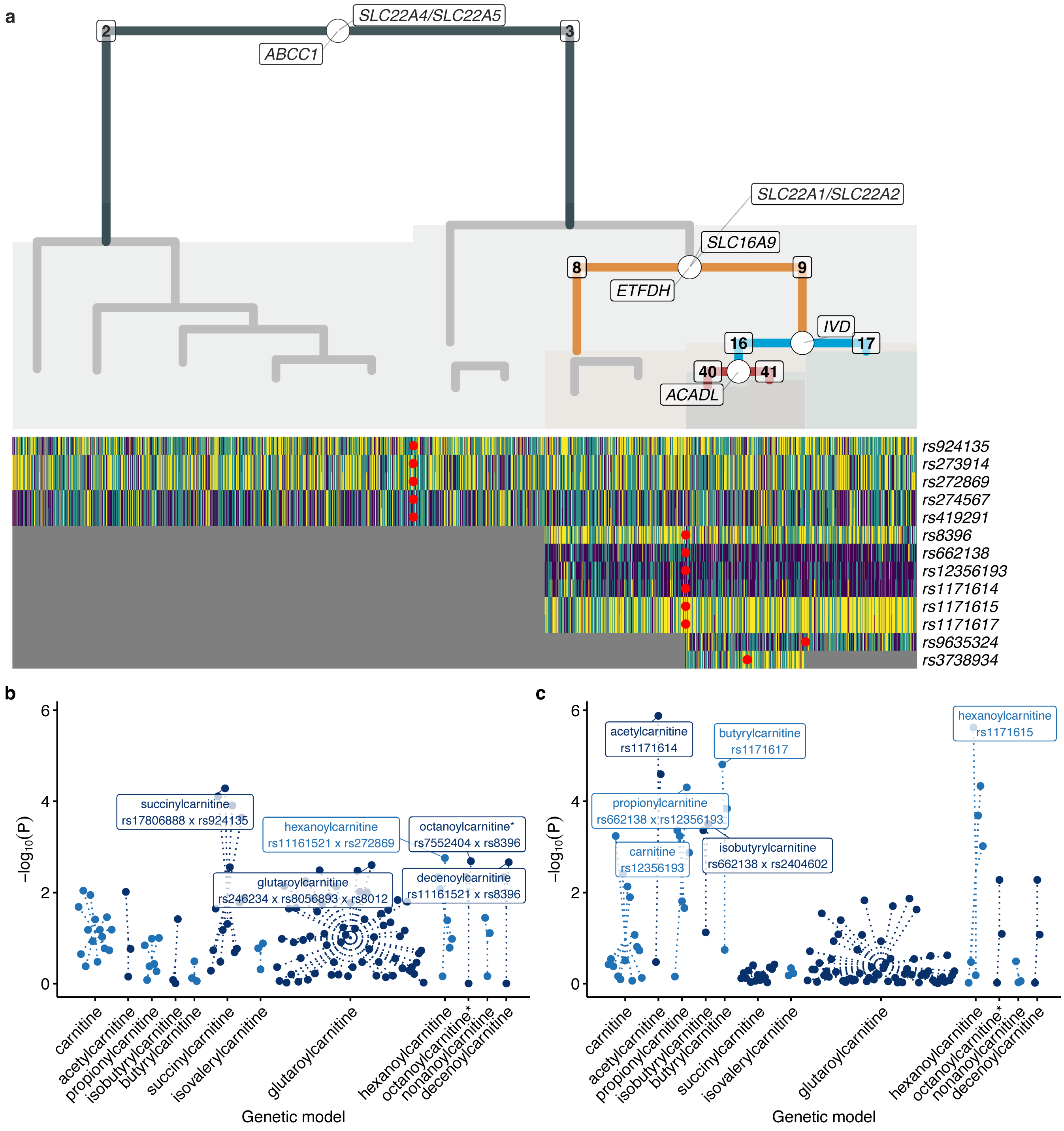

**Genetic associations with the acylcarnitine-derived clustering. (a)** Clustering from Figure 1a with additive associations with single acylcarnitine-associated SNPs and annotated genes in the dendrogram, respectively. Full association results are provided in Supplementary Table 7. Genotypic associations between both single SNPs and epistatic models with branching points were only significant for clusters 2 and 3 (**b**), and clusters 8 and 9 (**c**). Dotted lines connect genetic models for the same acylcarnitines in the Manhattan plots. Full association results are provided in Supplementary Table 9.

For the additive single-SNP analysis, we observed 13 significant ($P < 4.55 \times 10^{-3}$) SNP-cluster associations mapping to seven unique loci. Identified loci separating clusters 2 and 3 were *SLC22A4/SLC22A5* (4 SNPs; lead-SNP rs272869, $P = 2.24 \times 10^{-3}$) and *ABCC1* (rs924135, $P = 3.86 \times 10^{-5}$); *SLC16A9* (4 SNPs; lead-SNP rs1171614, $P = 3.04 \times 10^{-6}$), *SLC22A1/SLC22A2* (rs662138, $P = 4.28 \times 10^{-4}$) and *ETFDH* (rs8396, $P = 1.54 \times 10^{-3}$) separating clusters 8 and 9; *IVD* (rs9635324, $P = 3.53 \times 10^{-3}$) separating clusters 16 and 17; and *ACADL* (rs3738934, $P = 9.16 \times 10^{-4}$) separating clusters 40 and 41. Interestingly, all significant SNP associations were with cluster 3 or within its lower branches, suggesting that genetic contributions to the acylcarnitine clustering are more pronounced in subgroups with more progressed AD biomarker profiles.

The analysis of genetic interactions revealed no substantial improvements over single SNP analysis in terms of strength of association or explained variance. The notable exception was a 2-SNP epistatic model for succinylcarnitine (C4-DC) between rs17806888 (mapped to *SUCLG2*) and rs924135 (mapped to *ABCC1*). The association was only slightly more significant ($P = 5.19 \times 10^{-5}$; ANOVA test for interaction $P = 0.0218$) than the association of rs924135 alone ($P = 7.77 \times 10^{-5}$). However, the interaction model led to a boost in explained variance from 1.5% to over 30%. It has to be noted that analyses in lower branches of the dendrogram (i.e., below the branching points separating clusters 6 and 7 as well as cluster 8 and 9) were underpowered for this type of analysis, leading to large error estimates or non-convergence of more complex epistatic models.

### Supplementary Figure 3

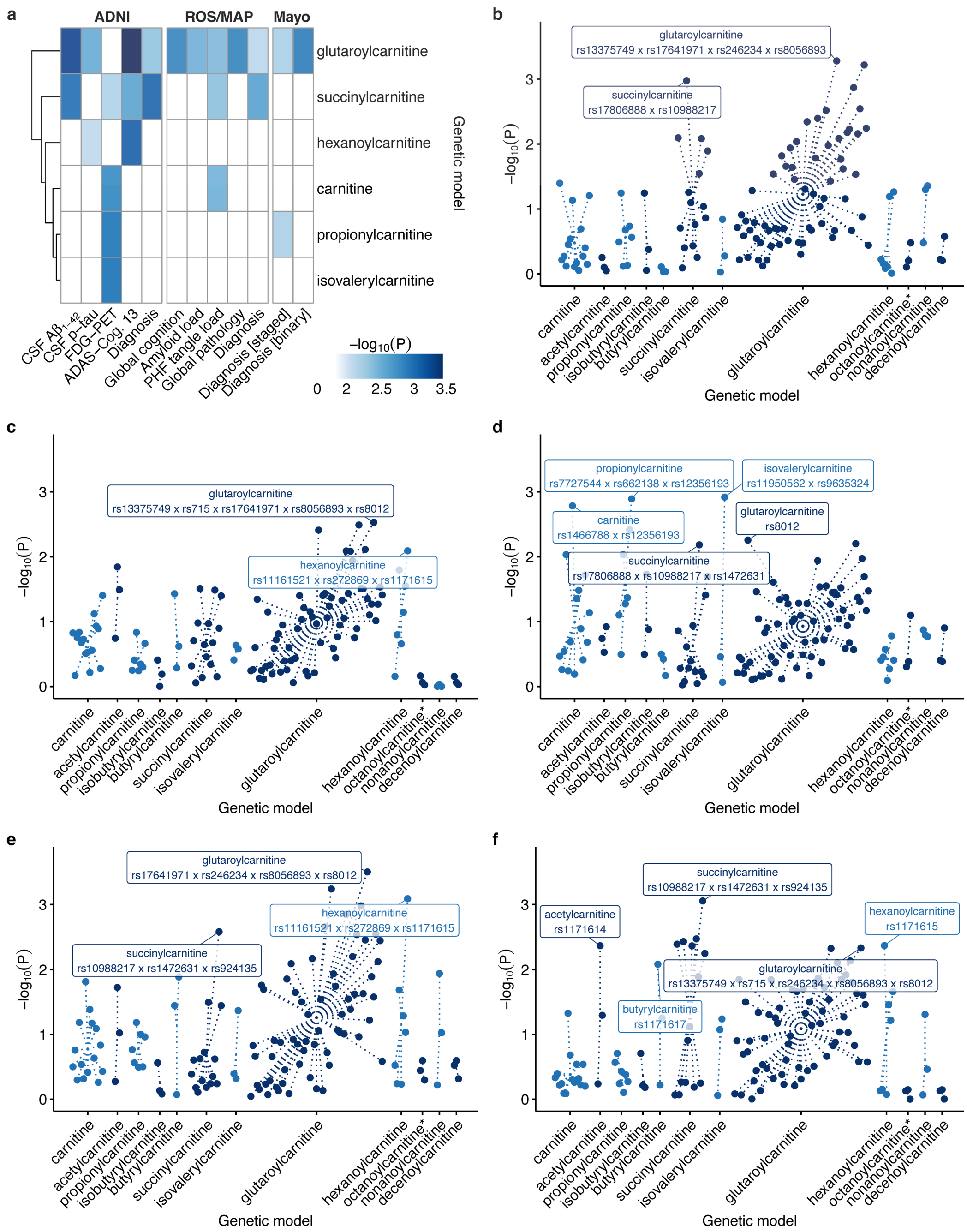

**Genetic associations of acylcarnitine-associated SNPs with A-T-(N)-(C) measures.** This figure presents a detailed view of single SNP and epistatic associations with A-T-(N)-(C) measures using genotypic tests. **(a)** Heatmap which condenses significant association results covering outcomes in ADNI, MayoLOAD, and ROS/MAP. Manhattan plots are included for (**b**) CSF $A\beta_{1\text{-}42}$ in ADNI, where dotted lines connect genetic models for the same acylcarnitines. Equivalent plots are provided for (**c**) CSF p-tau, (**d**) FDG-PET, (**e**) ADAS-Cog.13 , and (**f**) clinical diagnosis . Full results are provided in Supplementary Table 10. * The genetic models for octanoylcarnitine and decanoylcarnitine were identical, which is why we only show results for octanoylcarnitine.

This analysis revealed significant epistatic effects across all investigated clinical phenotypes in ADNI. Notably, these associations were limited to epistatic models for free carnitine (C0) and short-chain even and odd acylcarnitines (C2-C6), while no associations were found in epistatic analyses for acylcarnitines with longer chain lengths. Overall, the strongest interaction effect we observed was that of an epistatic model for glutarylcarnitine on CSF $A\beta_{1\text{-}42}$. The strongest association for CSF p-tau was again with an epistatic model for glutarylcarnitine; for FDG-PET was with the epistatic model for isovalerylcarnitine; for ADAS-Cog. 13 was with an epistatic model for glutarylcarnitine; and for diagnosis was with an epistatic model for succinylcarnitine. Of note, we only considered interaction effects to be informative if they were more significant than any association of the single SNPs contained in the respective epistatic model.

To strengthen confidence in our findings, we performed replication analysis of these epistatic associations with AD in two additional studies: ROS/MAP (n = 2,059) and MayoLOAD (n = 2,067), panel **(a)**. As in ADNI, both studies showed the strongest associations with AD or AD-linked pathological burden for epistatic models for glutarylcarnitine. The MayoLOAD study further replicated a significant epistatic association for propionylcarnitine, while the ROS/MAP study additionally replicated the epistatic associations for succinylcarnitine and free carnitine.

#### Supplementary Figure 4

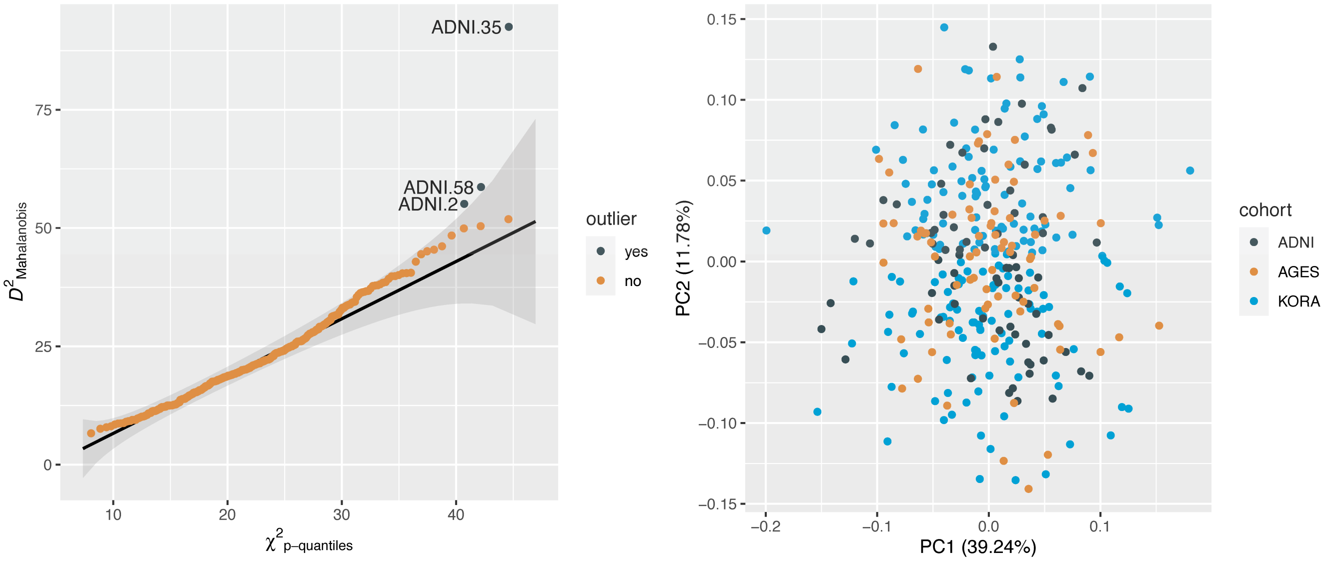

**Quality control of reference subject data used for cross-normalization.** The figure represents quality control assessments for reference subjects utilized for cohort cross-normalization in ADNI, AGES, and KORA. The left panel displays multivariable outlier detection using Mahalanobis distance, highlighting three ADNI reference subjects as outliers that were subsequently removed. The right panel presents a principal component analysis plot demonstrating no clear separation among reference subjects across cohorts.

#### Supplementary Figure 5

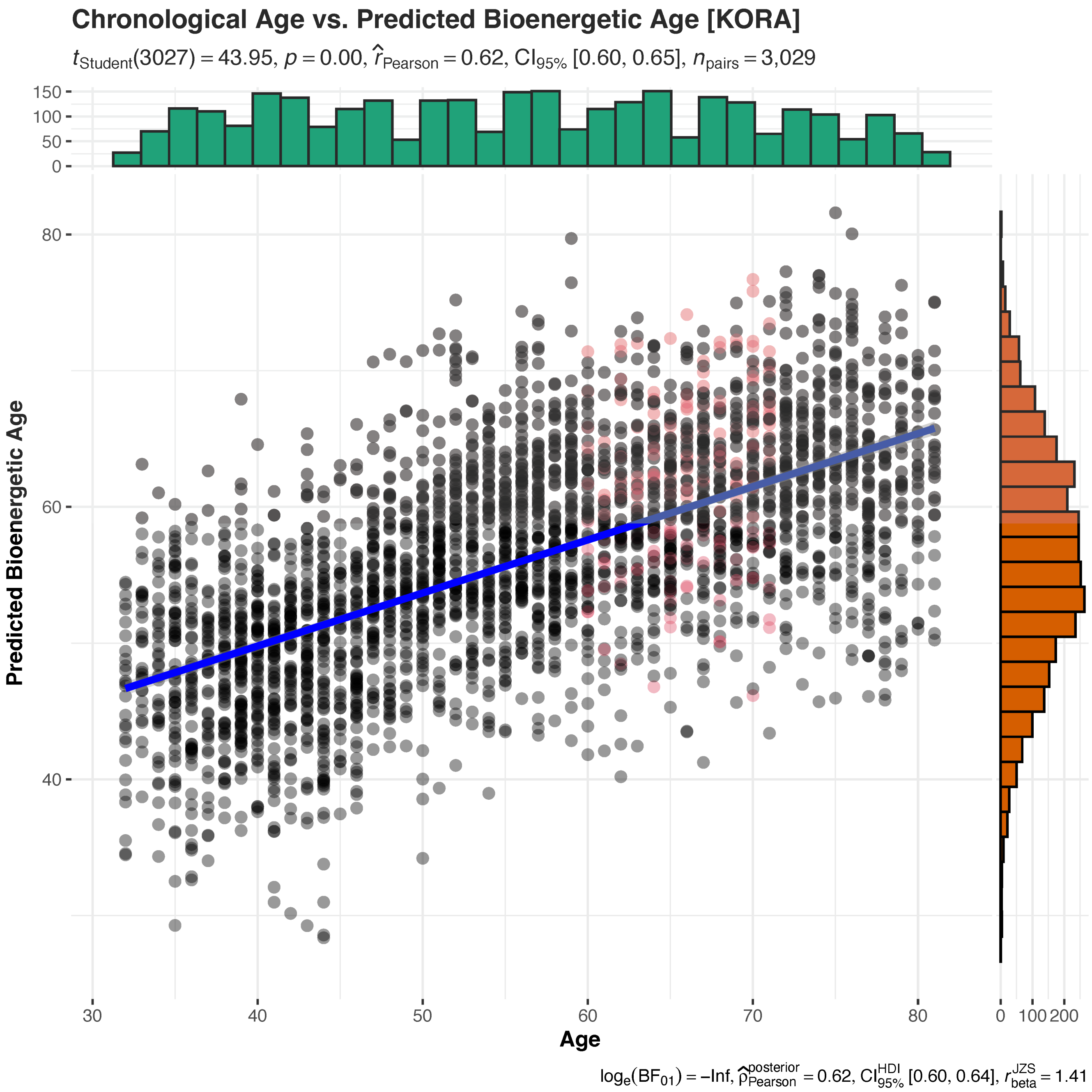

**Correlation plot of predicted bioenergetic age and chronological age in KORA.** This figure displays the correlation between the two age measures with statistics between predicted bioenergetic age and chronological age in KORA. Reference subjects used for cohort cross-normalization are highlighted in red. p=0.00 is shown because the p-value was smaller than 4.9e-324.

#### Supplementary Figure 6

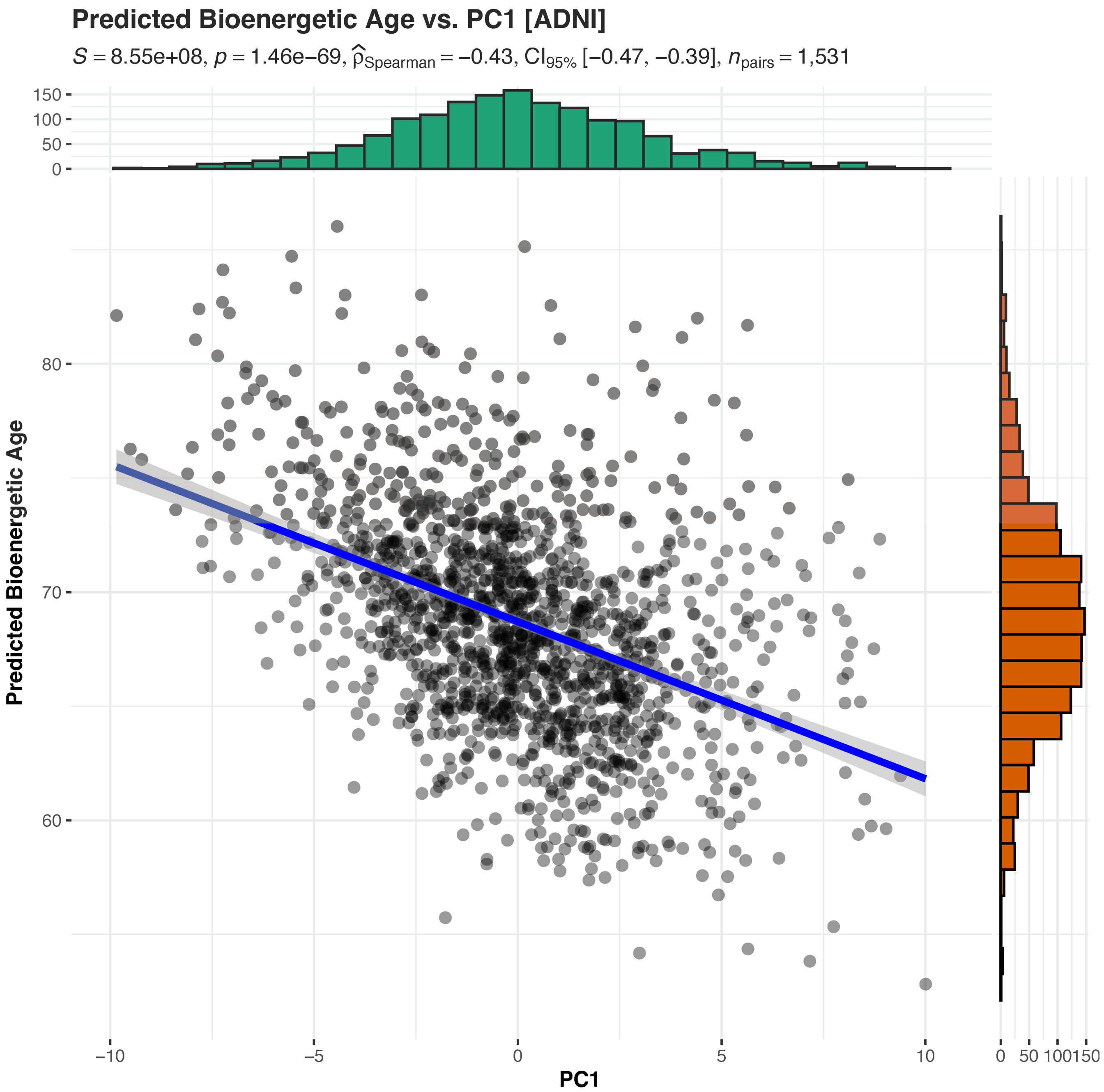

**Correlation plot of predicted bioenergetic age and Principal Component 1 of the acylcarnitine profiles underlying the clustering in ADNI.** The figure shows the strong correlation of the two measures, with statistics comparing the predicted bioenergetic age against the first principal component of acylcarnitine profiles of ADNI participants. The strong correlation indicates that the major clustering structure of the data is well represented by predicted bioenergetic age.

### Supplementary Figure 7

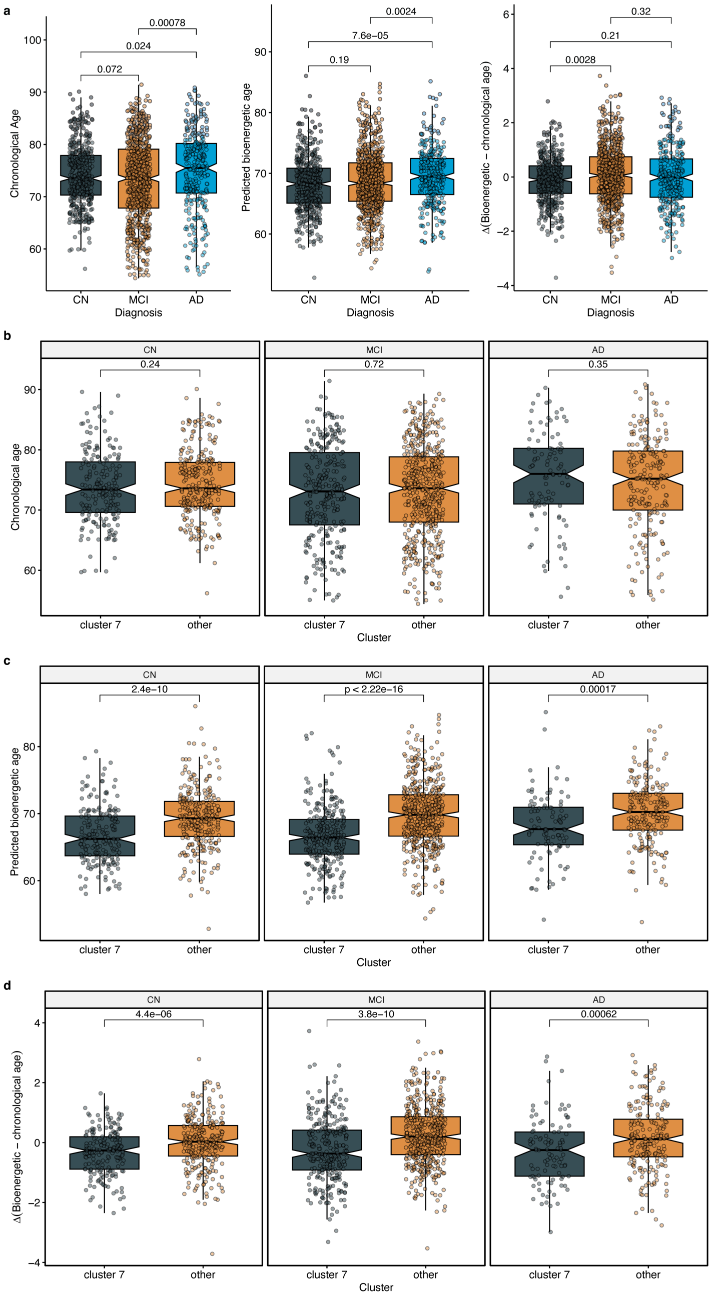

**Age measures across diagnostic groups for all ADNI participants and separately for those in cluster 7. (a)** Comparison of three age measures (chronological age, predicted bioenergetic age, and their delta) across diagnostic groups in the pooled sample. **(b-d)** Cluster 7 vs. all other ADNI participants, stratified by diagnostic groups.

This shows that the bioenergetic age effects in cluster 7 do not stem from the cluster consisting of younger individuals or a smaller fraction of individuals with symptoms. Rather, study participants in cluster 7 consistently present a chronological age that is comparable to the rest, yet their bioenergetic age is remarkably younger than that of other participants. This observation holds true across all diagnostic groups.

### Supplementary Figure 8

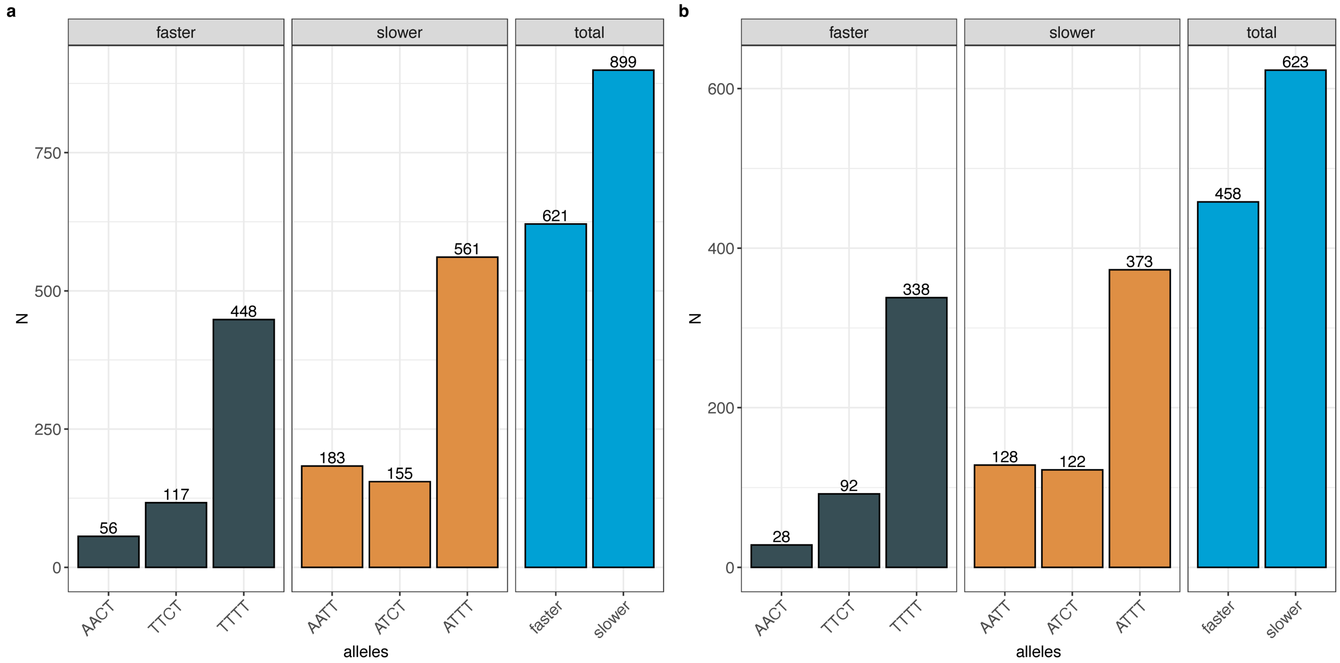

**Frequency of allele combinations in the binary grouping for slower/faster cognitive decline.** The frequency of allele combinations and their groupings were comparable between (**a**) ADNI and (**b**) ROS/MAP.
